# The Emerging Dominance of Genetic Disorders in Under-5 Mortality: A Global Comparative Assessment and Frontier Analysis, 1990–2021

**DOI:** 10.64898/2026.03.07.26347836

**Authors:** Jingjing Ruan, Zewen Tao, Kaiye Zhang, Shaoran Wu, Xiaoxiao Yu, Hao Zhang, Yue Zhang

**Affiliations:** Liangzhu Laboratory, Department of Genetics and Metabolism, Children’s Hospital, Zhejiang University School of Medicine, National Clinical Research Center for Children and Adolescents’ Health and Diseases, Hangzhou, Zhejiang 310000, China; Eye Center, The Second Affiliated Hospital, Zhejiang University School of Medicine, Hangzhou, Zhejiang 310009, China; Zhejiang University-University of Edinburgh Institute (ZJU-UoE Institute), Zhejiang University School of Medicine, International Campus, Zhejiang University, Haining, Zhejiang 314400, China

**Author notes:** Corresponding author: Yue Zhang. Jingjing Ruan, Zewen Tao and Kaiye Zhang contributed equally to this work.

**Keywords:** genetic disorders, under-5 mortality, Global Burden of Disease (GBD), congenital birth defects, frontier analysis

## Abstract

**Background:** Global under-5 mortality has declined by approximately 60% since 1990, driven largely by reductions in communicable, maternal, neonatal, and nutritional (CMNN) diseases. Yet the degree to which genetic disorders now impede further progress toward Sustainable Development Goal (SDG) 3.2 remains poorly quantified. No prior study has assessed the aggregate burden of genetically determined conditions as a unified category across the full spectrum of countries and development levels.

**Methods:** Using data from the Global Burden of Disease (GBD) Study 2021, we defined a composite “Total Genetic Burden” by aggregating 16 genetically determined causes of death, encompassing congenital birth defects, hemoglobinopathies, cystic fibrosis proxies, and spinal muscular atrophy proxies, across 204 countries and territories from 1990 to 2021. Age-standardized mortality rates (ASMR), proportional mortality ratios (PMR), years of life lost (YLLs), and 95% uncertainty intervals (UIs) were calculated. Temporal trends were assessed to evaluate the shifting burden over the study period. Age-specific mortality density was computed to identify periods of peak vulnerability. Deterministic frontier analysis (log-transformed quadratic quantile regression at the 5th percentile) was applied to quantify potentially avoidable mortality relative to best-observed global performance at each level of socioeconomic development.

**Results:** The age-standardized mortality rate of genetic disorders in children under 5 declined from 1990 to 2021; however, the proportional mortality ratio nearly doubled (from 5.76% to 10.76%), and genetic disorders rose from the fifth to the third leading cause of under-5 death. This shift was most pronounced in high Socio-demographic Index (SDI) countries, where genetic disorders accounted for over 40% of all under-5 deaths in some nations (e.g., Libya, 46.32%). An “Epidemiological Paradox” emerged: absolute mortality correlated negatively with SDI (R = −0.79, *P* < 0.001), whereas proportional mortality correlated positively (R = 0.80, *P* < 0.001). Age-specific analysis revealed a “Neonatal Stronghold,” with genetic disorders accounting for 57% of combined genetic-versus-infectious deaths in the first week of life but only 8% in children aged 1–4 years. Frontier analysis identified substantial efficiency gaps across all SDI quintiles; China and Japan sat on the effective frontier, while Afghanistan, Nigeria, and even the United States exhibited considerable potentially avoidable mortality.

**Conclusions:** Genetic disorders have shifted from a secondary concern to a leading structural barrier to further reductions in child mortality. Achieving SDG 3.2 will require broadening global child health priorities beyond infection control to include prenatal screening, newborn screening programs, and pediatric surgical capacity building, particularly in low- and middle-income countries.

## Background

Global under-5 mortality has fallen substantially over the past three decades. According to the United Nations Inter-agency Group for Child Mortality Estimation (UN IGME), the under-5 mortality rate declined from 93.0 per 1,000 live births in 1990 to 37.1 per 1,000 in 2021, a reduction of approximately 60%[1]. Much of this progress is attributable to the scale-up of interventions targeting communicable diseases: expanded immunization, oral rehydration therapy, insecticide-treated bed nets, and improved nutritional supplementation[2, 3]. As these threats have receded, however, the composition of residual under-5 mortality has changed. The causes of death that persist are increasingly those that resist conventional public health interventions and depend instead on complex healthcare infrastructure. Genetic disorders are chief among them[4, 5].

Despite this growing importance, the contribution of genetic disorders to global child mortality has been underestimated and fragmented in the literature. Prior studies have focused on individual disease categories, such as congenital heart defects[6], neural tube defects[7], or sickle cell disease[8], without capturing the aggregate burden of genetically determined conditions as a whole. The GBD classification system itself contributes to this fragmentation: congenital structural anomalies fall under “Congenital birth defects” at Level 2, hemoglobinopathies (including sickle cell disorders, thalassemias, and glucose-6-phosphate dehydrogenase [G6PD] deficiency) are categorized under hematological conditions, and monogenic diseases such as cystic fibrosis and spinal muscular atrophy (SMA) are scattered across respiratory and neurological disorder categories[9]. This taxonomic dispersion has prevented a comprehensive assessment of the total genetic burden on pediatric survival and has limited policymakers’ ability to recognize the full scope of the problem.

Analytically, existing burden-of-disease studies have largely relied on descriptive metrics such as average annual percentage change (AAPC) to characterize temporal trends, without using methods that can identify inflection points in disease trajectories or evaluate health system performance against empirical benchmarks[10, 11]. Frontier analysis, a technique increasingly used in global health research to quantify the gap between observed and best-achievable outcomes at a given level of socioeconomic development, has not been applied to pediatric genetic disorders[12, 13].

The present study had two objectives. First, we quantified the temporal shift in the relative importance of genetic disorders within under-5 mortality from 1990 to 2021 across 204 countries and territories, using a composite measure we term “Total Genetic Burden” that aggregates all genetically determined causes of death in the GBD framework. Second, we applied frontier analysis to identify countries with superior or inferior outcomes relative to their socioeconomic development level, thereby estimating the magnitude of potentially avoidable mortality attributable to health system inefficiencies rather than economic constraints.

## Methods

### Data source

Data were obtained from the Global Burden of Disease (GBD) Study 2021, coordinated by the Institute for Health Metrics and Evaluation (IHME) at the University of Washington[9]. The GBD 2021 provides estimates of mortality, morbidity, and disability for 371 diseases and injuries across 204 countries and territories, 21 GBD regions, and 7 super-regions from 1990 to 2021. Cause-of-death estimates were generated using the Cause of Death Ensemble model (CODEm), which synthesizes vital registration data, verbal autopsy surveys, cancer registries, and other epidemiological sources, with adjustments for data quality, completeness, and redistribution of garbage codes[14]. All estimates include 95% uncertainty intervals (UIs) derived from 1,000 posterior draws.

### Definition of “Total Genetic Burden”

We defined the “Total Genetic Burden” as a composite measure encompassing all GBD-classified causes of death in children under 5 years that are primarily determined by genetic or chromosomal abnormalities. This definition aggregated 16 specific cause categories distributed across multiple levels of the GBD cause hierarchy (Table 1). The core group consisted of nine categories within the GBD Level 2 classification of “Congenital birth defects”: congenital heart anomalies (Cause ID 641), neural tube defects (642), Down syndrome (643), other chromosomal abnormalities (644), congenital musculoskeletal and limb anomalies (645), urogenital congenital anomalies (646), digestive congenital anomalies (647), other congenital birth defects (648), and orofacial clefts (665). The extended group incorporated four hemoglobinopathy-related categories: sickle cell disorders (614), thalassemias (613), G6PD deficiency (615), and other hemoglobinopathies and hemolytic anemias (618). Additionally, two proxy categories were included to capture monogenic diseases not independently classified in the GBD framework: “Other chronic respiratory diseases” (Cause ID 520) as a proxy for cystic fibrosis, and “Motor neuron disease” (556) as a proxy for spinal muscular atrophy.

**Table 1.** Composition of “Total Genetic Burden”: included GBD cause categories.

| Category<br>(GBD Level 2/3) | Specific Cause Name<br>(GBD Level 3/4) | GBD<br>Cause ID | Inclusion Rationale |
| --- | --- | --- | --- |
| Congenital birth defects | Congenital heart anomalies | 641 | Core structural: leading cause of fatal birth defects globally |
| Congenital birth defects | Neural tube defects | 642 | Core structural: major neurological cause of neonatal death |
| Congenital birth defects | Down syndrome | 643 | Chromosomal: most common viable chromosomal disorder |
| Congenital birth defects | Other chromosomal abnormalities | 644 | Chromosomal: includes trisomy 13, 18, Turner, and Klinefelter syndromes |
| Congenital birth defects | Orofacial clefts | 665 | Structural: common, surgically correctable defect |
| Congenital birth defects | Digestive congenital anomalies | 647 | Structural: includes esophageal atresia, diaphragmatic hernia |
| Congenital birth defects | Urogenital congenital anomalies | 646 | Structural: includes renal agenesis and dysplasia |
| Congenital birth defects | Congenital musculoskeletal and limb anomalies | 645 | Structural: includes clubfoot and limb reduction defects |
| Congenital birth defects | Other congenital birth defects | 648 | Structural: unspecified structural malformations |
| Hemoglobinopathies | Sickle cell disorders | 614 | Functional (Blood): one of the most prevalent monogenic diseases globally |
| Hemoglobinopathies | Thalassemias | 613 | Functional (Blood): severe forms cause fatal anemia in early childhood |
| Hemoglobinopathies | G6PD deficiency | 615 | Functional (Enzyme): leading cause of kernicterus-related mortality |
| Hemoglobinopathies | Other hemoglobinopathies and hemolytic anemias | 618 | Functional (Blood): rare hemolytic conditions |
| Chronic respiratory diseases | Other chronic respiratory diseases | 520 | Functional (CF proxy): best available proxy for cystic fibrosis |
| Neurological disorders | Motor neuron disease | 556 | Functional (SMA proxy): best available proxy for spinal muscular atrophy |

Conditions of primarily environmental, infectious, or maternal etiology—including congenital syphilis, congenital Zika syndrome, fetal alcohol spectrum disorders, and complications of prematurity—were explicitly excluded to ensure the specificity of the genetic burden construct.

### Outcome metrics

Four primary metrics were extracted or calculated for the under-5 population (age 0–4 years) across all 204 countries and territories for each year from 1990 to 2021:

(1) Deaths: absolute number of deaths attributable to each included genetic cause, with 95% UIs. (2) Age-standardized mortality rate (ASMR): mortality rate per 100,000 population, standardized to the GBD reference population, enabling valid cross-national and temporal comparisons. (3) Years of life lost (YLLs): calculated as the sum of each death multiplied by the residual life expectancy at the age of death, based on a reference life table, reflecting the premature mortality impact of genetic disorders. (4) Proportional mortality ratio (PMR): defined as the number of deaths from genetic disorders divided by the total number of deaths from all causes in children under 5, expressed as a percentage. The PMR served as the primary metric for evaluating the relative importance of genetic disorders within the broader mortality landscape.

The Socio-demographic Index (SDI), a composite indicator ranging from 0 to 1 that incorporates per capita income, educational attainment, and total fertility rate, was used to stratify countries into five development quintiles (low, low-middle, middle, high-middle, and high SDI)[9].

### Statistical analysis

#### Frontier analysis

To evaluate health system performance independent of economic development, we conducted a deterministic frontier analysis following established methods applied in prior GBD publications[12, 13]. The Socio-demographic Index (SDI) served as the independent variable (x-axis), and the ASMR for genetic disorders served as the dependent variable (y-axis). The effective frontier curve was estimated using a log-transformed quadratic quantile regression at the 5th percentile, representing the empirical minimum mortality achievable at any given SDI level based on best-observed global performance. The “efficiency gap” for each country was defined as the vertical distance between its observed mortality rate and the corresponding point on the frontier curve, representing the magnitude of potentially avoidable mortality that could theoretically be eliminated through health system optimization without requiring changes in overall socioeconomic development[12].

#### Correlation and comparative analyses

Spearman rank correlation coefficients were used to assess the relationship between SDI and both the ASMR and PMR of genetic disorders at the country level. Age-specific vulnerability was evaluated by calculating mortality density (total annual deaths divided by the number of days in each standard pediatric age interval: early neonatal [0–6 days], late neonatal [7–27 days], post-neonatal [28–364 days], and early childhood [1–4 years]) to normalize for the vastly different durations of each age stratum. All analyses were conducted using R software (version 4.3.1; R Foundation for Statistical Computing, Vienna, Austria), with the packages ggplot2, sf, ggbump, frontier, and Benchmarking used for visualization and frontier estimation[15].

### Ethical considerations

This study used de-identified, publicly available, aggregate-level data from the GBD 2021 database. No individual-level data were accessed, and no ethical approval or informed consent was required.

## Results

### Spatial heterogeneity and the “Epidemiological Paradox”

In 2021, the burden of genetic disorders varied widely across 204 countries and territories (Figure 1). The absolute mortality rate (Figure 1A), depicted on a gradient from light pink (<50 per 100,000) to deep red (>200 per 100,000), was highest in low-SDI and low-middle-SDI regions. A belt of high burden stretched across sub-Saharan Africa, with the darkest shading concentrated in West Africa and the Horn of Africa, alongside parts of South Asia where Afghanistan and neighboring countries exceeded 200 per 100,000. Afghanistan had the highest age-standardized mortality rate at 219.2 per 100,000 (95% UI: 142.6–326.0), followed by Haiti (169.3 per 100,000) and Libya (151.4 per 100,000). Western Europe, East Asia, and Australasia had uniformly low absolute rates (<50 per 100,000), as shown in the inset maps.

**Figure 1.**
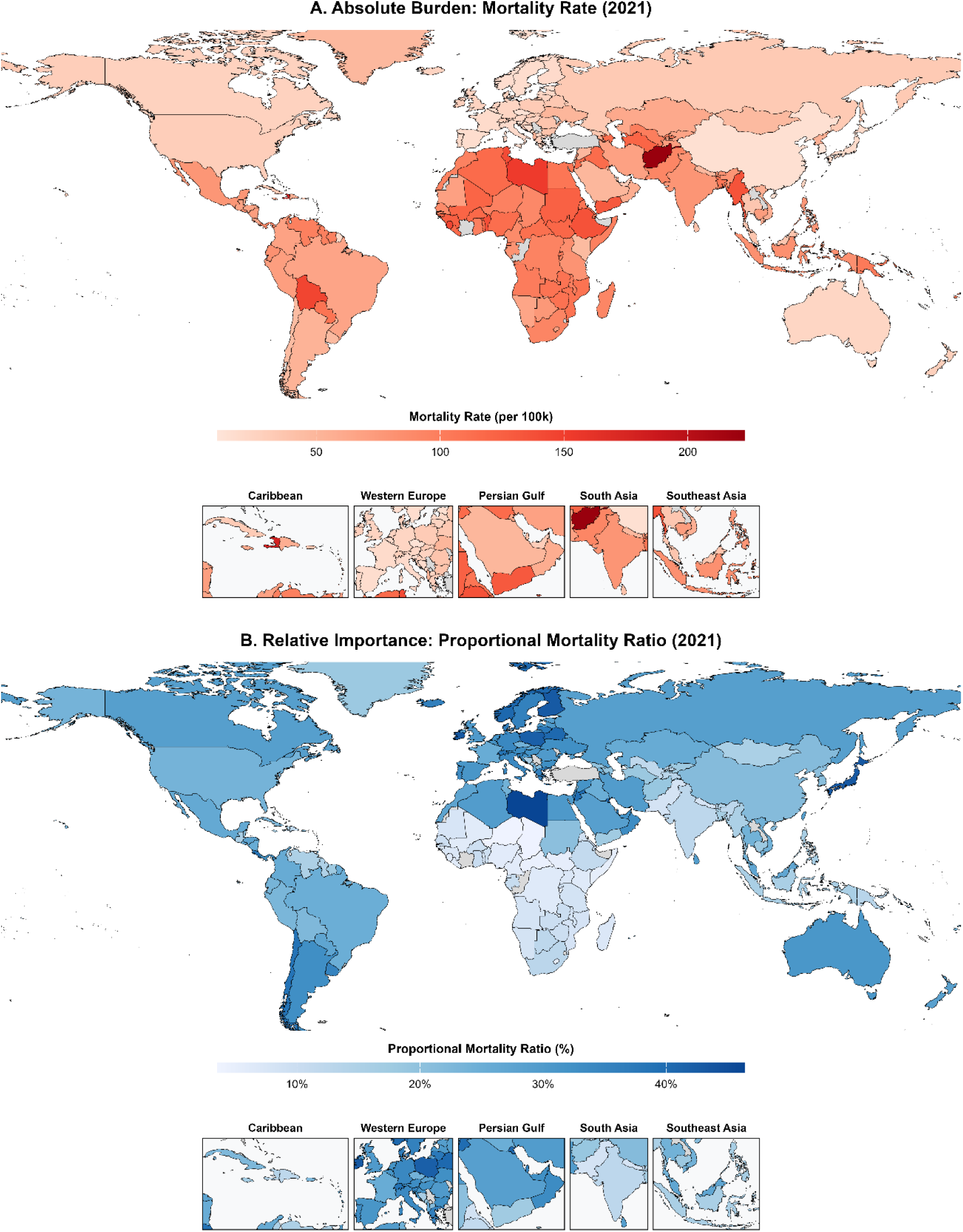
The global shift in burden of genetic disorders in children under 5, 2021. **(A)** Absolute burden: age-standardized mortality rate (per 100,000 population) of genetic disorders across 204 countries and territories. The color gradient ranges from light pink (low rates, <50 per 100,000) to deep red (high rates, >200 per 100,000), with the highest absolute burden concentrated in sub-Saharan Africa and South Asia. **(B)** Relative importance: proportional mortality ratio (PMR, %), representing the percentage of total under-5 deaths attributable to genetic disorders. The color gradient ranges from light blue (low proportions, <10%) to deep blue (high proportions, >40%), with the highest proportional burden observed in high-SDI regions including Western Europe, East Asia, and the Persian Gulf states. Inset maps provide sub-regional detail for the Caribbean, Western Europe, Persian Gulf, South Asia, and Southeast Asia. PMR, proportional mortality ratio.

A different picture emerged when the relative burden was examined (Figure 1B). The proportional mortality ratio (PMR), mapped from light blue (<10%) to deep blue (>40%), showed a geographic pattern that was largely inverted relative to the absolute burden map. Libya recorded the highest PMR globally at 46.32%, meaning that nearly half of all under-5 deaths in the country were attributable to genetic conditions. The Persian Gulf region, Western Europe, East Asia, and Australasia all had elevated PMRs (>25–40%). Sub-Saharan Africa, by contrast, displayed low PMRs (generally <15%) despite bearing the highest absolute mortality, because communicable diseases still dominate the total mortality denominator. This inversion constitutes what we term the “Epidemiological Paradox”: the countries with the lowest absolute genetic disorder mortality are the same countries where genetic disorders account for the largest share of child deaths.

This discordance between absolute burden and relative importance has a practical implication: in countries where the epidemiological transition is most advanced, genetic disorders are no longer a secondary concern but the primary barrier to further mortality reduction. The inset maps for the Caribbean, Western Europe, Persian Gulf, South Asia, and Southeast Asia reveal sub-regional variation obscured in the global view, including high PMRs in small island states and Gulf nations with advanced healthcare systems.

### Temporal trends and rising prominence in mortality rankings

From 1990 to 2021, the composition of pediatric mortality shifted steadily, with communicable and genetic causes of death diverging in opposite directions (Figure 2A). The proportional contribution of communicable, maternal, neonatal, and nutritional (CMNN) diseases to total under-5 mortality fell from approximately 85% in 1990 to approximately 77% in 2021 (orange line). The PMR of genetic disorders rose in parallel, from 5.76% to 10.76% over the same period (blue line). The two trend lines did not intersect, as CMNN diseases still account for most child deaths, but the widening gap between them captures a gradual rebalancing: as CMNN diseases recede, genetic disorders fill an increasing share of the residual mortality.

**Figure 2.**
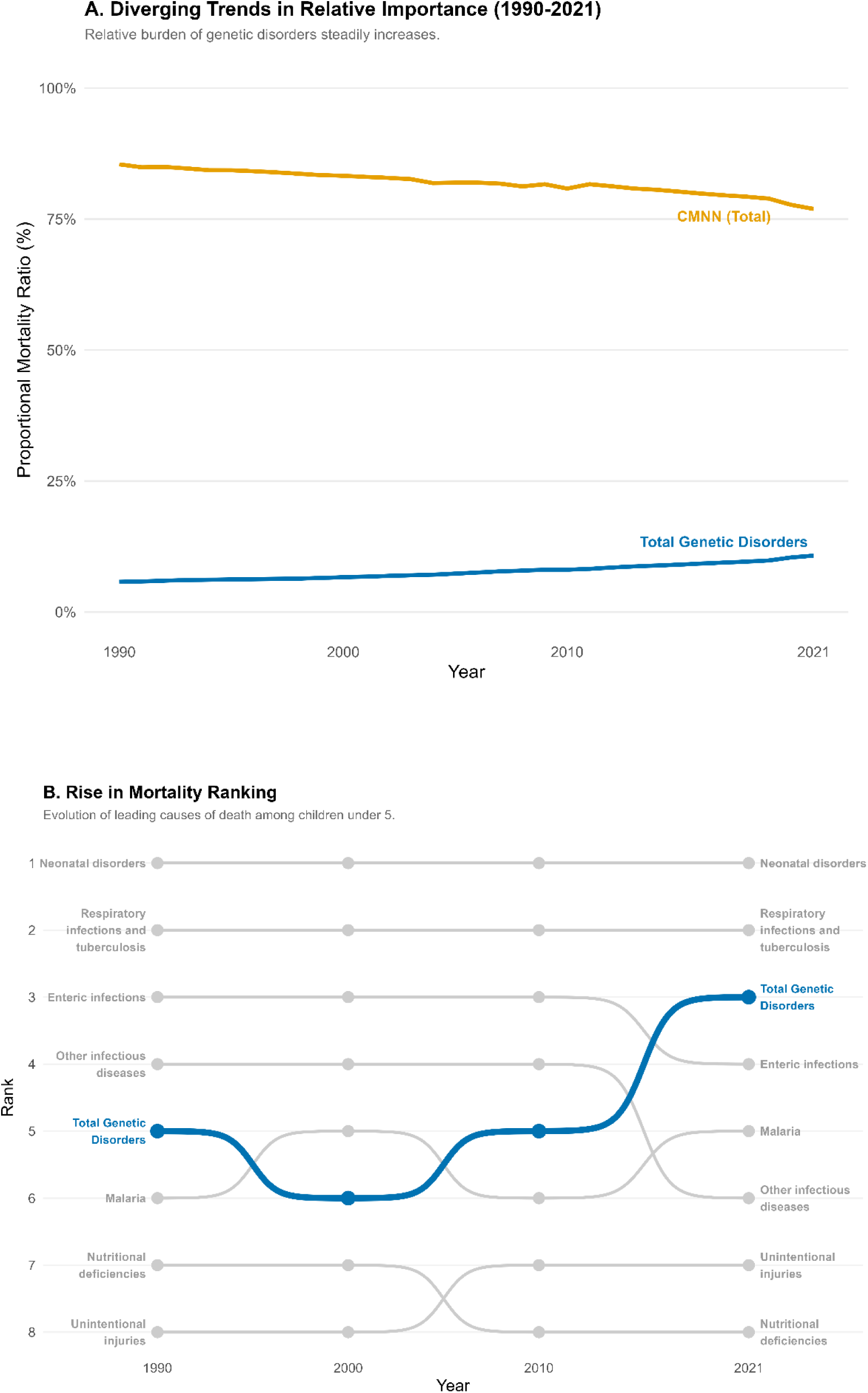
Diverging trends in relative importance and rise in mortality ranking for genetic disorders, 1990–2021. **(A)** Diverging trends in relative importance (1990–2021): proportional mortality ratio (PMR, %) for total genetic disorders (blue line) versus communicable, maternal, neonatal, and nutritional (CMNN) diseases (orange line). The relative burden of genetic disorders steadily increases while the CMNN share gradually declines, illustrating a progressive divergence pattern. **(B)** Rise in mortality ranking: bump chart depicting the evolution of leading causes of death among children under 5 at four time points (1990, 2000, 2010, 2021). Total genetic disorders (highlighted in blue) remain at rank 5 from 1990 to 2010 before ascending sharply to rank 3 by 2021, surpassing enteric infections and other infectious diseases. Neonatal disorders and respiratory infections maintain stable positions at ranks 1 and 2, respectively. CMNN, communicable, maternal, neonatal, and nutritional diseases; PMR, proportional mortality ratio.

This divergence was accompanied by a rise in the cause-of-death ranking (Figure 2B). The bump chart shows that genetic disorders held steady at the 5th leading cause of under-5 death from 1990 through 2010, behind neonatal disorders (1st), respiratory infections and tuberculosis (2nd), enteric infections (3rd–4th), and other infectious diseases (which exchanged positions in the middle ranks). Between 2010 and 2021, however, genetic disorders surpassed both enteric infections and other infectious diseases to become the 3rd leading cause of death. This rank shift in the final decade contrasts with two decades of stability and coincides with the period of most rapid decline in diarrheal disease mortality, driven by rotavirus vaccination and improved sanitation. While the overall global age-standardized mortality rate (ASMR) for genetic disorders declined during the study period, the pace of this reduction has noticeably slowed since 2010. This deceleration suggests diminishing returns from existing public health interventions when applied to complex congenital conditions.

This ascending rank does not mean genetic disorders are becoming more common in absolute terms. Rather, the rise reflects a differential pace of mortality reduction: CMNN diseases have benefited from highly cost-effective, scalable interventions (vaccines, antibiotics, oral rehydration therapy), while genetic disorders, which require complex diagnostics, surgical capacity, and long-term management, have seen comparatively modest reductions. They therefore account for a progressively larger share of the deaths that remain. The persistence of neonatal disorders at rank 1 throughout the study period reinforces this interpretation, as many neonatal deaths share the same dependence on advanced healthcare infrastructure.

### Association between socioeconomic development and genetic disorder burden

The relationship between socioeconomic development and genetic disorder burden ran in opposite directions depending on the metric used (Figure 3). SDI correlated negatively with the absolute mortality rate (R = −0.79, P < 0.001; Figure 3A). The LOESS regression curve was non-linear: mortality rates plateaued at approximately 110 per 100,000 among low-SDI countries (SDI 0.15–0.40), declined steeply through the middle-SDI range, and leveled off at approximately 10–30 per 100,000 among high-SDI nations. Countries in the lowest SDI quintile, including Afghanistan (approximately 225 per 100,000), Haiti (approximately 175 per 100,000), Sierra Leone, Benin, and Ethiopia, had the highest absolute mortality, a pattern consistent with limited healthcare infrastructure, scarce surgical capacity, absent prenatal screening, and co-existing infectious disease burdens that worsen outcomes for children with genetic conditions. Two outliers among higher-SDI countries stood out: Libya (SDI approximately 0.72) and Tunisia (SDI approximately 0.68) had mortality rates of approximately 150 and 115 per 100,000, respectively, far above the values predicted by their SDI and well outside the LOESS confidence band. Bolivia (Plurinational State of) similarly had higher-than-expected mortality for its SDI level.

**Figure 3.**
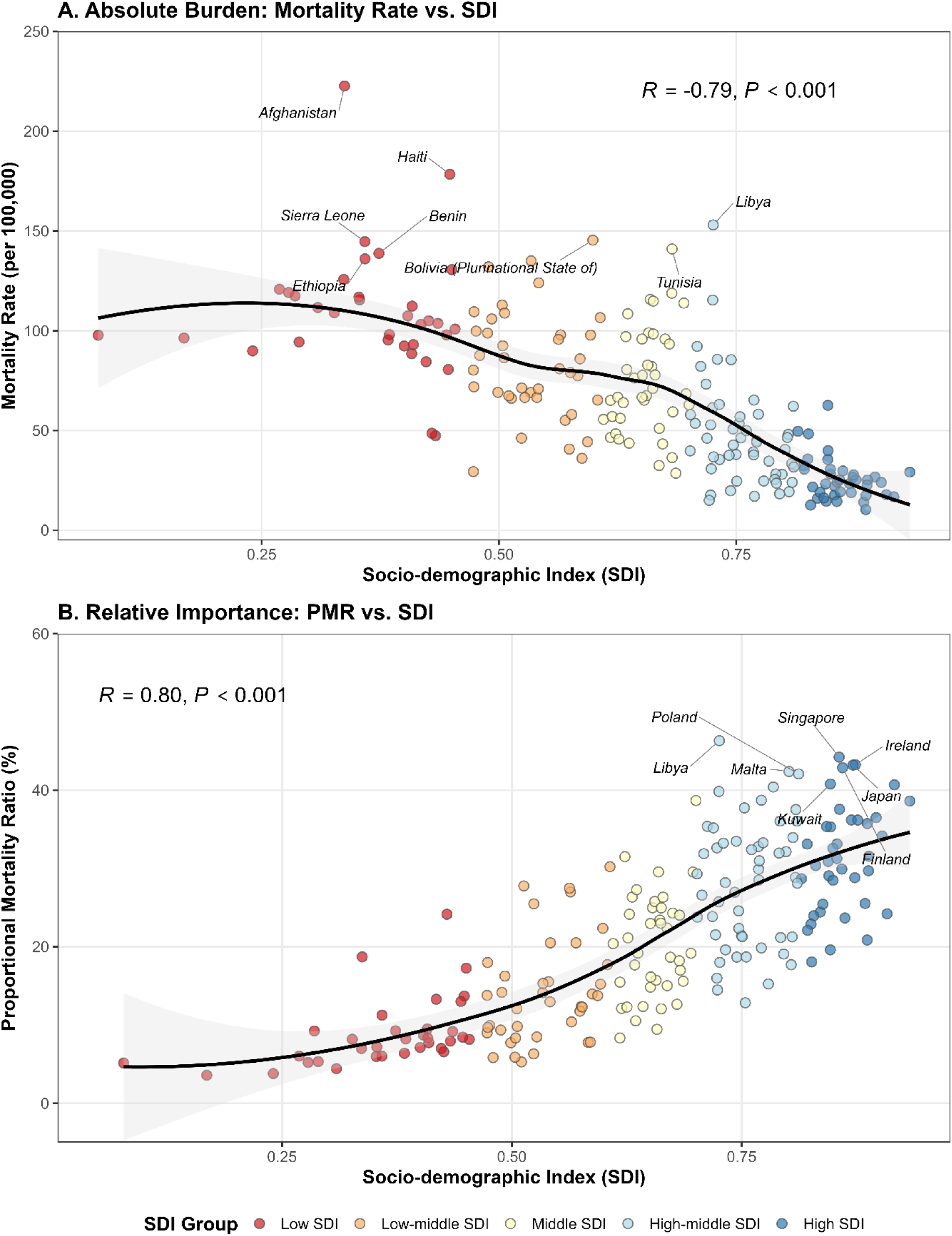
Correlation between the Socio-demographic Index (SDI) and genetic disorder burden across 204 countries and territories, 2021. **(A)** Absolute burden: mortality rate (per 100,000 population) versus SDI. The LOESS regression curve (solid black line) with 95% confidence interval (gray shading) demonstrates a non-linear negative relationship, with a plateau phase at low SDI followed by steep decline. Labeled countries include Afghanistan, Haiti, Sierra Leone, Benin, Ethiopia, and Bolivia (high absolute burden) as well as Libya and Tunisia (outliers with unexpectedly high mortality relative to their SDI). **(B)** Relative importance: proportional mortality ratio (PMR, %) versus SDI. The LOESS curve shows a consistent positive relationship. Labeled countries include Poland, Libya, Malta, Singapore, Kuwait, Finland, Japan, and Ireland, all exhibiting elevated PMRs above the regression trend. Data points are color-coded by SDI quintile (red: Low SDI; orange: Low-middle SDI; yellow: Middle SDI; light blue: High-middle SDI; dark blue: High SDI). Correlation coefficients (R) and P values are displayed. PMR, proportional mortality ratio; SDI, Socio-demographic Index.

The relationship between SDI and the proportional mortality ratio (PMR) was positive (R = 0.80, P < 0.001; Figure 3B). As countries controlled communicable causes of child death, genetic disorders accounted for a progressively larger proportion of the deaths that remained. The LOESS curve rose from approximately 4% at the lowest SDI values to approximately 30–35% at the highest. Several high or high-middle SDI countries had PMRs well above the regression trend: Singapore (approximately 48%), Ireland (approximately 48%), Poland (approximately 45%), Libya (approximately 42%), Malta (approximately 42%), and Japan (approximately 40%) all clustered above the LOESS curve. Kuwait and Finland were also notable. The wide dispersion of data points above the regression line in the high-SDI range indicates that even among the most developed countries, the degree to which genetic disorders dominate residual mortality varies considerably, likely reflecting differences in consanguinity rates, population genetic structure, screening infrastructure, and tertiary neonatal care quality. The scatter of country-level data around the regression curves in both panels was itself informative: countries at similar SDI levels often had markedly different genetic disorder mortality rates. Libya, for example, appeared as an outlier in both panels (high mortality and high PMR), suggesting that country-specific factors beyond aggregate socioeconomic development play a decisive role. This observation motivated the frontier analysis, which formally quantifies these performance differentials.

### Compositional shifts and the epidemiological transition within genetic disorders

The internal composition of the Total Genetic Burden differed by level of socioeconomic development (Figure 4). At the global level, congenital birth defects (green) consistently accounted for over 90% of total genetic disease mortality across all four time points (1990, 2000, 2010, 2021; Figure 4A). This dominance was essentially unchanged over 32 years, with only marginal shifts in the non-structural components visible in the lower portion of the split axis. The residual fraction, comprising sickle cell disorders (orange), other hemoglobinopathies and hemolytic anemias (blue), thalassemias (pink), G6PD deficiency (bright pink), other chronic respiratory diseases (yellow, cystic fibrosis proxy), and rare systemic conditions (purple, SMA proxy), collectively accounted for less than 15% of the global total. This “denominator effect” masked the dynamics of these less prevalent but clinically important conditions at the aggregate level.

**Figure 4.**
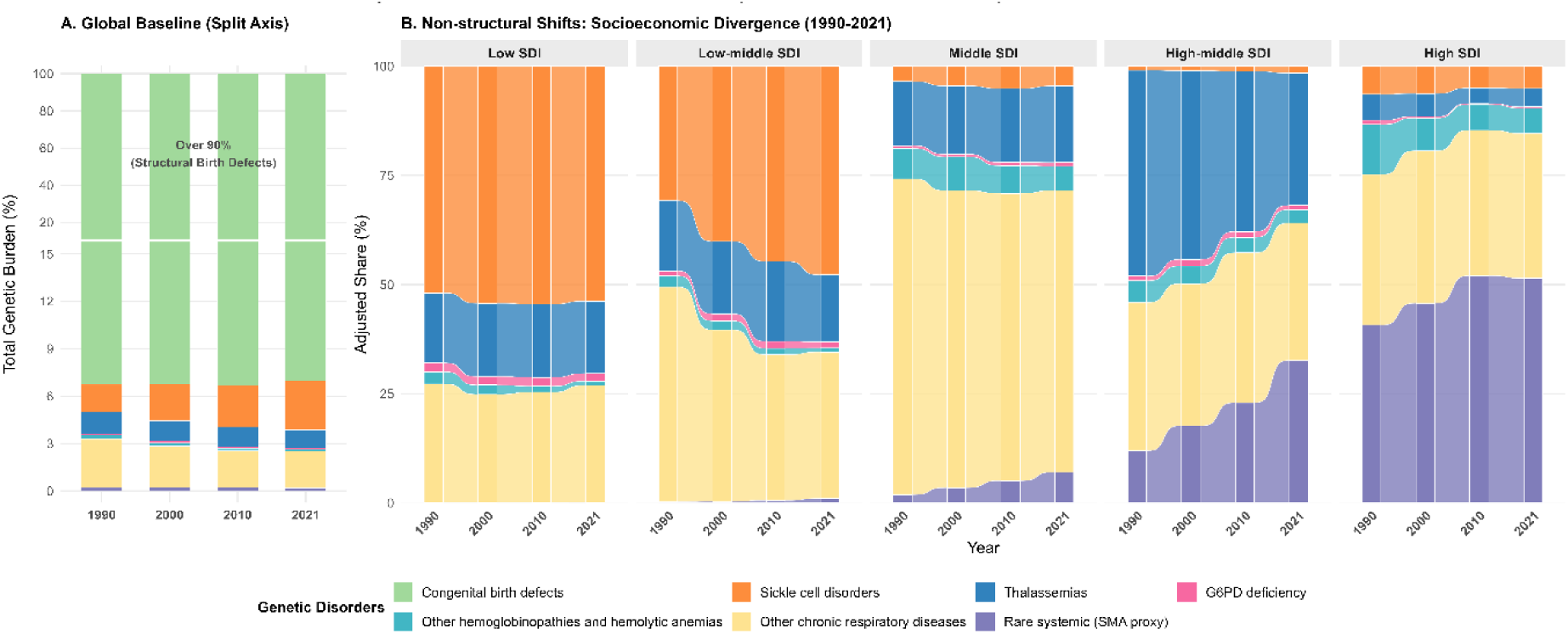
Structural and compositional dynamics of genetic mortality across SDI quintiles, 1990–2021. **(A)** Global baseline composition (split axis): stacked bar chart with a split y-axis showing the total genetic burden composition at four time points (1990, 2000, 2010, 2021). Congenital birth defects (green) consistently account for over 90% of total genetic mortality. The lower portion of the split axis expands the non-structural components, including sickle cell disorders (orange), other hemoglobinopathies and hemolytic anemias (blue), thalassemias (pink), G6PD deficiency (bright pink), other chronic respiratory diseases (yellow), and rare systemic/SMA proxy (purple). **(B)** Non-structural shifts and socioeconomic divergence (1990–2021): five stacked area charts showing the adjusted proportional share (%) of non-structural genetic disorders within each SDI quintile over time, with structural birth defects excluded. In low and low-middle SDI settings, endemic hemoglobinopathies (sickle cell disorders and thalassemias) dominate with minimal temporal change. In high-middle and high SDI settings, a marked compositional transition is evident, with rare systemic conditions (SMA proxy, purple) and other chronic respiratory diseases (CF proxy, yellow) progressively displacing hemoglobinopathies to become the predominant non-structural contributors by 2021. SDI, Socio-demographic Index; SMA, spinal muscular atrophy; CF, cystic fibrosis.

To reveal the compositional transitions hidden beneath this dominance, we excluded structural birth defects and examined the adjusted proportional shares of the remaining non-structural genetic disorders across five SDI quintiles separately (Figure 4B). The stratified analysis exposed a clear epidemiological divergence by socioeconomic context.

In the low-SDI quintile, the non-structural mortality composition was stable from 1990 to 2021, dominated by sickle cell disorders (orange, approximately 50–55% of the adjusted share) and other hemoglobinopathies and hemolytic anemias (blue, approximately 20–25%), with smaller contributions from thalassemias and G6PD deficiency. The low-middle SDI quintile displayed a similar pattern of compositional stasis, though with a somewhat larger thalassemia component. In both quintiles, virtually no temporal change was observed over the 32-year period, indicating a persistent and unmitigated burden of endemic hemoglobinopathies in settings where malaria selective pressure maintains high carrier frequencies and where disease-modifying interventions such as hydroxyurea, chronic transfusion programs, and hematopoietic stem cell transplantation remain largely unavailable.

The middle-SDI quintile represented a transitional zone, displaying a more heterogeneous composition with growing contributions from thalassemias, other chronic respiratory diseases (yellow), and the early emergence of rare systemic conditions (purple), alongside a gradual reduction in the relative share of sickle cell disorders.

The largest compositional changes occurred in the high-middle and high SDI quintiles. In the high-middle SDI quintile, the other chronic respiratory diseases category (yellow, cystic fibrosis proxy) and rare systemic conditions (purple, SMA proxy) expanded progressively, collectively displacing hemoglobinopathies as the leading non-structural contributors by 2021. In the high-SDI quintile, this shift was most pronounced: rare systemic conditions grew from a minor fraction in 1990 to approximately 40–50% of the adjusted non-structural share by 2021, while other chronic respiratory diseases also occupied a substantial and growing proportion. Sickle cell disorders and other hemoglobinopathies were reduced to minimal slivers. This shift reflects a transition toward a disease profile dominated by conditions that require advanced diagnostics, expensive pharmacotherapies (e.g., gene therapy, enzyme replacement therapy, nusinersen), and multidisciplinary chronic care, resources concentrated in higher-income healthcare systems.

### Age-specific vulnerability and the “Neonatal Stronghold”

To account for the different durations of standard pediatric age intervals, we calculated mortality density, defined as total annual deaths divided by the number of days in each age interval, to capture the intensity of mortality risk per unit of time (Figure 5A). This analysis, presented as a back-to-back pyramid chart on a square-root scale, revealed a pronounced “Neonatal Stronghold” for genetic disorders. In 2021, the mortality density for genetic conditions peaked at 24,282 deaths per day during the early neonatal period (0–6 days), exceeding all other age strata by an order of magnitude. The density fell to 2,662 deaths per day in the late neonatal period (7–27 days), 615 in the post-neonatal period (28 days–1 year), and 54 in early childhood (1–4 years), a 450-fold reduction from the first week to early childhood. Common infections and nutritional deficiencies showed a different pattern: their early neonatal mortality density was also high (18,670 deaths per day), but the subsequent decline was less steep (4,758 in the late neonatal period, 2,468 post-neonatal, and 634 in early childhood), indicating sustained attrition beyond the neonatal period. The crossover between the two categories occurred between the early and late neonatal periods: genetic disorders exceeded infectious diseases in the first week (24,282 versus 18,670) but were surpassed from the late neonatal period onward (2,662 versus 4,758).

**Figure 5.**
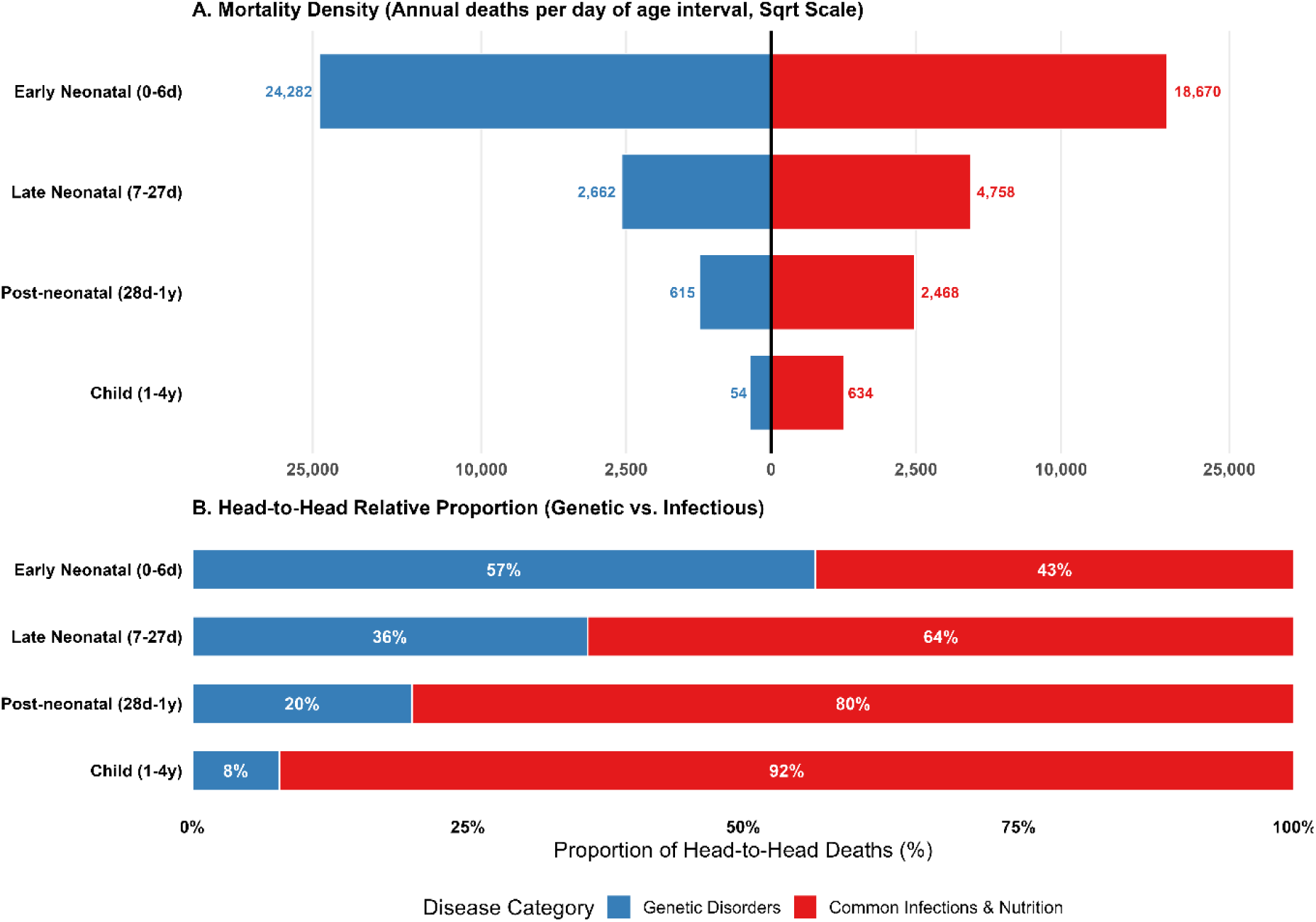
Age-specific vulnerability and the “Neonatal Stronghold,” 2021. **(A)** Mortality density (annual deaths per day of age interval) for genetic disorders (blue bars, extending left) versus common infections and nutritional deficiencies (red bars, extending right) across four pediatric age strata, presented as a back-to-back pyramid chart. A square-root scale is applied to the horizontal axis to accommodate the large magnitude differences between age groups. Specific mortality density values are labeled: early neonatal (0–6 days): 24,282 (genetic) versus 18,670 (infectious); late neonatal (7–27 days): 2,662 versus 4,758; post-neonatal (28 days–1 year): 615 versus 2,468; early childhood (1–4 years): 54 versus 634. **(B)** Head-to-head relative proportion: 100% stacked horizontal bar chart illustrating the shifting proportion of deaths between genetic disorders (blue) and common infections and nutrition (red) within each age stratum. Genetic disorders account for 57% of combined deaths in the early neonatal period, declining to 36%, 20%, and 8% in subsequent age groups.

The relative proportion between genetic and infectious causes across age strata reinforced this pattern (Figure 5B). Genetic disorders accounted for 57% of the combined deaths from these two categories during the first week of life (versus 43% for infections and nutrition). This proportion declined with age: 36% in the late neonatal period (versus 64%), 20% post-neonatal (versus 80%), and only 8% in early childhood (versus 92%), an almost complete reversal of mortality dominance within a single year of life.

These temporal dynamics have direct clinical and policy implications. The concentration of genetic disorder mortality in the first days of life, with a per-day death rate 450 times higher in the early neonatal period than in early childhood, defines a narrow intervention window. Conditions such as critical congenital heart defects, severe chromosomal abnormalities (trisomy 13/18), and inborn errors of metabolism exert their lethal effects within hours to days of birth; post-hoc interventions are largely ineffective unless preceded by prenatal detection or immediately available neonatal intensive care. That genetic disorders outnumber infectious deaths on a per-day basis during the first week of life, counter to the conventional assumption that infections dominate neonatal mortality, argues for expanded prenatal screening (including non-invasive prenatal testing and fetal echocardiography) and universal newborn screening (NBS) with rapid turnaround times as core components of strategies to reduce genetic disorder mortality.

### Frontier analysis: quantifying the global “efficiency gap”

Although the correlational analyses established an inverse relationship between SDI and genetic disorder mortality, the frontier analysis revealed wide heterogeneity in health system performance among countries at similar development levels (Figure 6). The effective frontier curve, estimated via log-transformed quadratic quantile regression at the 5th percentile (solid green line), represents the best-observed outcomes at each SDI level, declining from approximately 55 per 100,000 at the lowest SDI values (approximately 0.10) to approximately 5 per 100,000 at the highest (approximately 0.90). The vertical distance between a country’s observed mortality rate and the frontier defines the “efficiency gap,” the portion of mortality that is potentially avoidable through health system improvement rather than economic growth.

**Figure 6.**
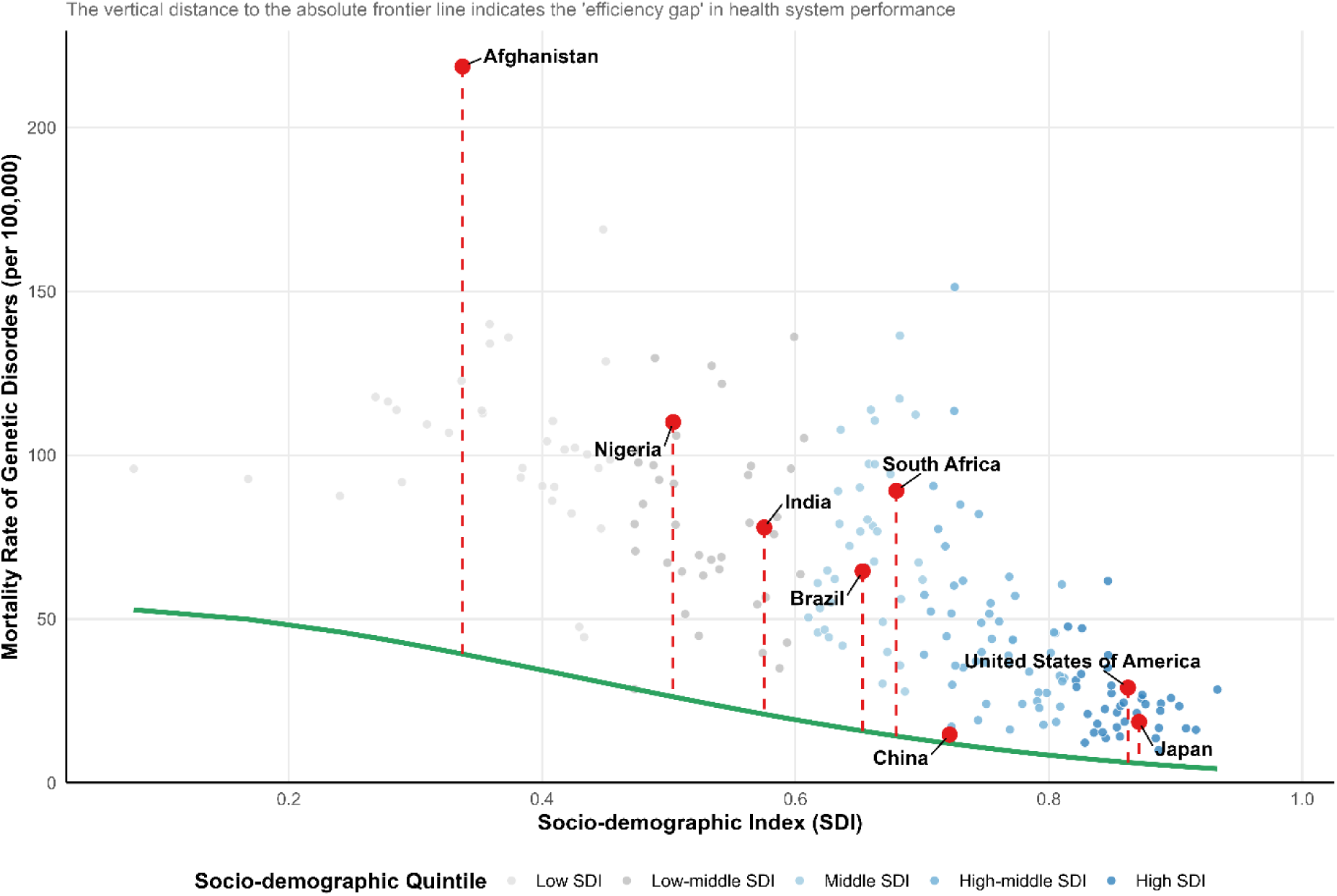
Frontier analysis of potentially avoidable genetic disorder mortality, 2021. Scatter plot displaying the relationship between the Socio-demographic Index (SDI, x-axis, range 0.1–1.0) and the mortality rate of genetic disorders (per 100,000 population, y-axis, range 0–220+) across 204 countries and territories. Background data points are color-coded by SDI quintile (gray: Low SDI; light gray: Low-middle SDI; light blue: Middle SDI; medium blue: High-middle SDI; dark blue: High SDI). The solid green curve represents the deterministic effective frontier, estimated via log-transformed quadratic quantile regression at the 5th percentile, declining from approximately 55 per 100,000 at the lowest SDI to approximately 5 per 100,000 at the highest SDI. Eight representative countries are highlighted with large red dots and labeled with dashed red vertical lines indicating their efficiency gaps: Afghanistan (SDI 0.30, mortality 220), Nigeria (SDI 0.48, mortality 110), India (SDI 0.55, mortality 80), Brazil (SDI 0.63, mortality 65), South Africa (SDI 0.70, mortality 95), China (SDI 0.72, mortality 15), United States of America (SDI 0.87, mortality 28), and Japan (SDI 0.92, mortality 18). China is positioned on the frontier curve, representing optimal performance at its development level. The vertical distance from each country to the frontier quantifies the potentially avoidable mortality achievable through health system optimization. SDI, Socio-demographic Index.

In the low and low-middle SDI tiers, Afghanistan (SDI approximately 0.30, mortality approximately 220 per 100,000) exhibited the largest efficiency gap of any country globally, with an observed mortality rate exceeding the frontier value at its SDI level (approximately 40 per 100,000) by nearly 180 per 100,000. Nigeria (SDI approximately 0.48, mortality approximately 110 per 100,000) similarly demonstrated a massive gap above the frontier (approximately 30 per 100,000 at its SDI level). These gaps likely reflect deficiencies in basic healthcare infrastructure (inadequate surgical capacity for congenital heart defects, absence of folic acid fortification for neural tube defect prevention, lack of newborn screening) rather than an inherent inability to reduce mortality at their development level.

Among middle and high-middle SDI countries, the frontier analysis showed large disparities in the translation of economic resources into health outcomes. India (SDI approximately 0.55, mortality approximately 80 per 100,000) and Brazil (SDI approximately 0.63, mortality approximately 65 per 100,000) both had substantial gaps above the frontier. South Africa (SDI approximately 0.70, mortality approximately 95 per 100,000) was the most prominent outlier among higher-SDI nations, with observed mortality approximately 75 per 100,000 above the frontier at its development level (approximately 20 per 100,000). Its mortality was comparable to countries at far lower SDI levels, suggesting that fragmented healthcare delivery, urban-rural disparities in specialist access, and the burden of the HIV/AIDS epidemic on pediatric health systems impede the translation of resources into better genetic disease outcomes. The efficiency gap persisted even in the United States of America (SDI approximately 0.87, mortality approximately 28 per 100,000), which had a mortality rate approximately 20 per 100,000 above the frontier (approximately 8 per 100,000 at its SDI level), indicating that high national income does not automatically produce optimal genetic disorder outcomes.

China (SDI approximately 0.72, mortality approximately 15 per 100,000) sat directly on the frontier curve, achieving the minimum mortality predicted for its development level. This performance likely reflects nationwide newborn screening, rapid expansion of pediatric cardiac surgical capacity, and integration of prenatal genetic screening into routine maternal care. Japan (SDI approximately 0.92, mortality approximately 18 per 100,000) was also near the frontier, consistent with its long-established investment in neonatal intensive care and universal screening.

The frontier analysis shows that socioeconomic development is necessary but not sufficient for reducing genetic disorder mortality. The extent of potentially avoidable mortality, even in countries with substantial economic resources such as the United States and South Africa, supports the case for health system optimization, targeted investment in genetic services, and cross-national learning from frontier-performing countries such as China and Japan.

## Discussion

### Principal findings in context

This study provides the first globally comparative assessment of genetic disorders as a unified category within under-5 mortality, spanning 204 countries and territories over 32 years. The findings show that while the absolute burden of genetic disorders has declined, their relative importance has nearly doubled, with their ranking rising from fifth to third leading cause of under-5 death between 1990 and 2021. This shift, which we term the “Emerging Dominance” of genetic disorders, does not reflect an increase in incidence but rather a differential pace of mortality reduction. The successful scale-up of interventions against communicable diseases has exposed a residual burden of genetically determined mortality that is resistant to conventional public health approaches.

These findings are consistent with the epidemiological transition framework proposed by Omran [16] and refined by later scholars [17]. The classical model predicts that as societies develop, the dominant causes of mortality shift from infectious and nutritional diseases to chronic and degenerative conditions. Our data show that this transition is operative within the pediatric age group, with genetic disorders acting as the primary “post-transition” threat to child survival. The transition is not uniform: high-SDI nations have already completed this shift, with genetic disorders accounting for over 40% of under-5 deaths in some cases, while low-SDI countries remain in the early stages, where the genetic burden is superimposed on a persistently high infectious disease burden rather than replacing it.

### The “Hidden Epidemic”: mechanisms of underestimation

The genetic disorder burden reported here is likely a conservative estimate. In many low-income countries with limited vital registration, a substantial proportion of neonatal deaths, particularly those in the first 24 hours, are classified as “neonatal sepsis,” “birth asphyxia,” or “unknown cause” without diagnostic workup for genetic conditions [18]. Limited availability of genetic testing, cytogenetic analysis, and post-mortem examination in these settings compounds the problem, producing systematic ascertainment bias that underestimates genetically determined mortality. The GBD modeling framework itself depends on input data quality: in countries without congenital anomaly surveillance systems, estimates rely more on regional covariates and predictive modeling than on direct observation, adding further uncertainty [9].

Additionally, our proxy-based approach for capturing cystic fibrosis (via “Other chronic respiratory diseases”) and spinal muscular atrophy (via “Motor neuron disease”) is an acknowledged limitation, as these aggregate categories encompass non-genetic conditions as well. These proxies were selected as the best available approximations within the GBD classification but may introduce misclassification in both directions. Sensitivity analyses excluding them did not materially alter the principal findings.

### Drivers of inequality: from diagnostics to surgical access

The efficiency gaps identified through frontier analysis reflect several health system deficiencies that disproportionately affect the management of genetic disorders relative to communicable diseases.

First, the diagnostic gap. Effective management of genetic conditions depends on timely detection, ideally prenatally or within the first days of life, yet the availability of prenatal screening (including NIPT and fetal anomaly ultrasound), newborn metabolic screening, and neonatal pulse oximetry for critical congenital heart defects varies enormously across countries [19]. High-income countries have expanded their newborn screening panels to 30–50 or more conditions; many low- and middle-income countries lack any systematic screening at all, resulting in delayed diagnosis and preventable deaths [20].

Second, the surgical capacity gap. Structural birth defects account for over 90% of the total genetic mortality burden, and congenital heart anomalies, the single largest contributor, require access to pediatric cardiac surgery, which remains concentrated in high-income countries. The Lancet Commission on Global Surgery estimated that five billion people lack access to safe, affordable surgical care [21]. The large efficiency gaps in countries such as Afghanistan and Nigeria are consistent with the documented absence of pediatric cardiac surgical capacity in these settings.

Third, the chronic care gap. Sickle cell disease is manageable with early diagnosis through newborn screening, penicillin prophylaxis, hydroxyurea, and comprehensive care programs, yet mortality from sickle cell disease in sub-Saharan Africa remains high because most affected countries lack these basic interventions [8, 22].

### Policy implications

These findings argue for broadening global child health priorities beyond the infection-focused paradigm that has guided pediatric survival efforts for the past three decades. Several policy directions follow from the data.

First, universal newborn screening (NBS) should be recognized as a core component of essential health services. The “Neonatal Stronghold” finding, showing that genetic disorders exert their greatest mortality impact in the first week of life, provides an epidemiological rationale for NBS programs with rapid turnaround times, even in resource-constrained settings where the initial panel may be limited to a few high-impact conditions (e.g., sickle cell disease, congenital hypothyroidism, critical congenital heart defects via pulse oximetry) [20, 23].

Second, the frontier analysis provides a framework for identifying countries where investment in genetic services is most likely to yield mortality reductions. Countries with large efficiency gaps at middle-SDI levels, where basic healthcare infrastructure exists but specialized genetic services do not, are high-yield targets for capacity building: regional referral centers for pediatric surgery, genetic counseling services, and prenatal diagnosis programs.

Third, the compositional analysis across SDI quintiles demonstrates that a “one-size-fits-all” approach to genetic disorder prevention is inappropriate. In low-SDI settings where hemoglobinopathies dominate the non-structural genetic burden, the most cost-effective interventions include carrier screening programs, folic acid food fortification for neural tube defect prevention, and integration of sickle cell disease management into existing maternal and child health platforms. In high-SDI settings where rare monogenic diseases constitute a growing share of genetic mortality, the policy focus should shift toward expanding access to orphan drug therapies, gene therapy, and advanced neonatal intensive care.

### Strengths and limitations

This study has several strengths. To our knowledge, it is the first to define and quantify a “Total Genetic Burden” by aggregating all genetically determined causes of under-5 mortality within the GBD framework, overcoming the fragmentation of prior single-disease analyses. The application of frontier analysis to pediatric genetic disorders is new and provides policy-relevant insights beyond descriptive epidemiology. The 32-year temporal scope and 204-country coverage allow identification of both global trends and country-specific patterns that narrower analyses would miss.

Several limitations must be acknowledged. First, the accuracy of GBD estimates varies substantially across countries, with estimates for nations lacking comprehensive vital registration systems relying more heavily on statistical modeling and covariates. This may affect the precision of country-specific estimates, particularly in sub-Saharan Africa and parts of South Asia. Second, the GBD classification system does not permit the isolation of certain monogenic diseases (e.g., cystic fibrosis, SMA) as independent causes, necessitating the use of proxy categories that may introduce misclassification. Third, the ecological nature of this analysis precludes causal inference at the individual level; the associations between SDI and genetic disorder burden should be interpreted as population-level correlations rather than causal relationships. Fourth, the COVID-19 pandemic may have introduced transient distortions in mortality data for 2020–2021, potentially affecting the most recent year of our analysis through disruptions to healthcare access, data collection systems, and vital registration processes [24]. Fifth, the GBD does not provide clinical severity grading for genetic conditions, precluding the distinction between inherently lethal conditions (e.g., trisomy 13/18) and potentially treatable conditions (e.g., isolated congenital heart defects), which limits the precision of estimates of “avoidable” mortality.

## Conclusions

This analysis of 204 countries over 32 years shows that genetic disorders have risen in relative importance within global under-5 mortality. Absolute mortality rates have declined, but the proportional mortality ratio has nearly doubled, and genetic disorders are now the third leading cause of death in children under 5. The “Epidemiological Paradox,” in which the highest absolute mortality occurs in the poorest countries while the highest relative burden is in the most developed, reveals a challenge that cuts across income levels. Frontier analysis confirms that considerable mortality is potentially avoidable through health system improvement at every level of socioeconomic development.

Without broader recognition that genetic disorders now constitute a leading structural barrier to reducing child mortality, and without corresponding investment in newborn screening, prenatal diagnostics, pediatric surgical capacity, and disease-modifying therapies, Sustainable Development Goal 3.2 (reducing under-5 mortality to below 25 per 1,000 live births by 2030) will remain out of reach in many countries. Child survival strategies must expand to include genetic service delivery as a routine component of pediatric healthcare.

## Acknowledgement

We thank the collaborators of the Global Burden of Diseases, Injuries, and Risk Factors Study 2021 for their work in generating and maintaining these estimates, and the Institute for Health Metrics and Evaluation for making the GBD data publicly accessible.

## Authors’ contributions

J.R., Z.T., and K.Z. contributed equally to this work, share first authorship, and conceptualized and designed the study. Z.T. curated the data, developed the methodology, and performed the statistical analyses. K.Z. and S.W. contributed to data curation and interpretation of the findings. J.R. drafted the original manuscript. X.Y., H.Z., and Y.Z. critically revised and reviewed the manuscript. All authors read and approved the final manuscript.

## Data availability

All data used in this study are publicly available from the Global Health Data Exchange (GHDx) at https://ghdx.healthdata.org/gbd-2021.

## Ethical approval

Not required. This study used publicly available, de-identified, aggregate data from the GBD 2021 database.

## References

1. United Nations Inter-agency Group for Child Mortality Estimation (UN IGME). Levels & Trends in Child Mortality: Report 2022. New York: United Nations Children’s Fund (UNICEF); 2023.

2. Liu L, Oza S, Hogan D, Chu Y, Perin J, Zhu J, et al. Global, regional, and national causes of under-5 mortality in 2000-15: an updated systematic analysis with implications for the Sustainable Development Goals. Lancet. 2016;388:3027–35. 10.1016/S0140-6736(16)31593-8.

3. Perin J, Mulick A, Yeung D, Villavicencio F, Lopez G, Strong KL, et al. Global, regional, and national causes of under-5 mortality in 2000-19: an updated systematic analysis with implications for the Sustainable Development Goals. Lancet Child Adolesc Health. 2022;6:106–15. 10.1016/S2352-4642(21)00311-4.

4. Modell B, Berry RJ, Boyle CA, Christianson A, Darlison M, Dolk H, et al. Global regional and national causes of child mortality. Lancet. 2012;380:1556; author reply 1556-1557. 10.1016/S0140-6736(12)61878-9.

5. Christianson, Arnold, Howson, Christopher P., Modell, Bernadette. March of Dimes Global Report on Birth Defects: The Hidden Toll of Dying and Disabled Children. White Plains, NY: March of Dimes Birth Defects Foundation; 2026.

6. GBD 2017 Congenital Heart Disease Collaborators. Global, regional, and national burden of congenital heart disease, 1990-2017: a systematic analysis for the Global Burden of Disease Study 2017. Lancet Child Adolesc Health. 2020;4:185–200. 10.1016/S2352-4642(19)30402-X.

7. Blencowe H, Kancherla V, Moorthie S, Darlison MW, Modell B. Estimates of global and regional prevalence of neural tube defects for 2015: a systematic analysis. Ann N Y Acad Sci. 2018;1414:31–46. 10.1111/nyas.13548.

8. Piel FB, Steinberg MH, Rees DC. Sickle Cell Disease. N Engl J Med. 2017;376:1561–73. 10.1056/NEJMra1510865.

9. GBD 2021 Diseases and Injuries Collaborators. Global incidence, prevalence, years lived with disability (YLDs), disability-adjusted life-years (DALYs), and healthy life expectancy (HALE) for 371 diseases and injuries in 204 countries and territories and 811 subnational locations, 1990-2021: a systematic analysis for the Global Burden of Disease Study 2021. Lancet. 2024;403:2133–61. 10.1016/S0140-6736(24)00757-8.

10. Kim HJ, Fay MP, Feuer EJ, Midthune DN. Permutation tests for joinpoint regression with applications to cancer rates. Stat Med. 2000;19:335–51. 10.1002/(sici)1097-0258(20000215)19:3%3C335::aid-sim336%3E3.0.co;2-z.

11. Clegg LX, Hankey BF, Tiwari R, Feuer EJ, Edwards BK. Estimating average annual per cent change in trend analysis. Stat Med. 2009;28:3670–82. 10.1002/sim.3733.

12. GBD 2015 Healthcare Access and Quality Collaborators. Electronic address, GBD 2015 Healthcare Access and Quality Collaborators. Healthcare Access and Quality Index based on mortality from causes amenable to personal health care in 195 countries and territories, 1990-2015: a novel analysis from the Global Burden of Disease Study 2015. Lancet. 2017;390:231–66. 10.1016/S0140-6736(17)30818-8.

13. GBD 2016 Healthcare Access and Quality Collaborators. Measuring performance on the Healthcare Access and Quality Index for 195 countries and territories and selected subnational locations: a systematic analysis from the Global Burden of Disease Study 2016. Lancet. 2018;391:2236–71. 10.1016/S0140-6736(18)30994-2.

14. Foreman KJ, Lozano R, Lopez AD, Murray CJ. Modeling causes of death: an integrated approach using CODEm. Popul Health Metr. 2012;10:1. 10.1186/1478-7954-10-1.

15. R Core Team. R: A Language and Environment for Statistical Computing. 2023.

16. Omran AR. The epidemiologic transition. A theory of the epidemiology of population change. Milbank Mem Fund Q. 1971;49:509–38.

17. Gaziano JM. Fifth phase of the epidemiologic transition: the age of obesity and inactivity. JAMA. 2010;303:275–6. 10.1001/jama.2009.2025.

18. Lawn JE, Blencowe H, Oza S, You D, Lee ACC, Waiswa P, et al. Every Newborn: progress, priorities, and potential beyond survival. Lancet. 2014;384:189–205. 10.1016/S0140-6736(14)60496-7.

19. Therrell BL, Padilla CD, Loeber JG, Kneisser I, Saadallah A, Borrajo GJC, et al. Current status of newborn screening worldwide: 2015. Semin Perinatol. 2015;39:171–87. 10.1053/j.semperi.2015.03.002.

20. Modell B, Darlison M. Global epidemiology of haemoglobin disorders and derived service indicators. Bull World Health Organ. 2008;86:480–7. 10.2471/blt.06.036673.

21. Meara JG, Leather AJM, Hagander L, Alkire BC, Alonso N, Ameh EA, et al. Global Surgery 2030: evidence and solutions for achieving health, welfare, and economic development. Lancet. 2015;386:569–624. 10.1016/S0140-6736(15)60160-X.

22. Grosse SD, Odame I, Atrash HK, Amendah DD, Piel FB, Williams TN. Sickle cell disease in Africa: a neglected cause of early childhood mortality. Am J Prev Med. 2011;41 6 Suppl 4:S398–405. 10.1016/j.amepre.2011.09.013.

23. Wilson, James Maxwell Glover, Jungner, Gunnar. Principles and Practice of Screening for Disease. Geneva: World Health Organization; 1968.

24. GBD 2021 Causes of Death Collaborators. Global burden of 288 causes of death and life expectancy decomposition in 204 countries and territories and 811 subnational locations, 1990-2021: a systematic analysis for the Global Burden of Disease Study 2021. Lancet. 2024;403:2100–32. 10.1016/S0140-6736(24)00367-2.

